# Psychosocial Stress and Allostatic Load Among Underrepresented Minority Women with Familial Cancer Risk

**DOI:** 10.64898/2026.08.26.26361226

**Authors:** Eliya K. Shachar, Roni Haas, Victoria E. Rodriguez, Jenny Lester, Mehrnaz A. Siavoshi, Lorna Kwan, Mariana Niell-Swiller, Paul Spellman, Paul C. Boutros, Vivian Y. Chang, Beth Y. Karlan

**Author notes:** Co-first author (Eliya K. Shachar, Roni Haas). **Correspondence**: Dr. Beth Y. Karlan, UCLA Jonsson Comprehensive Cancer Center David Geffen School of Medicine at UCLA, CHS-27-117.

## Abstract

**Importance:** Chronic stress may contribute to adverse health outcomes through cumulative physiologic dysregulation. Allostatic load (AL), a composite measure of multisystem physiologic burden, may capture biologic effects of structural, social, and psychosocial stress not reflected by self-reported measures.

**Objective:** To evaluate racial and ethnic differences in AL among women with familial cancer risk and examine how socioeconomic status, psychosocial factors, clinical characteristics, and health behaviors contribute to variations in AL.

**Design:** Cross-sectional study of underrepresented minority participants enrolled in the HERSTORY cohort from October 2023 through September 2025, with comparison participants from the UCLA ATLAS biobank.

**Setting:** UCLA academic health system.

**Participants:** The study included 303 racially and ethnically diverse female HERSTORY participants aged ≥35 years with a family history of cancer and matched non-Hispanic White female ATLAS participants (n=709).

**Exposures:** Race and ethnicity, age, neighborhood deprivation, cancer history and stage, depression, perceived stress, cancer worry, and physical activity.

**Main Outcomes and Measures:** The primary outcome was AL, calculated from cardiometabolic and organ-function measures. A secondary index incorporated race– and ethnicity-specific neutrophil-to-lymphocyte ratio (NLR) derived from 326,826 women in the UCLA Health population. Multivariable regression models evaluated factors associated with elevated AL.

**Results:** Compared with matched non-Hispanic White participants, Black and Asian/Pacific Islander HERSTORY participants had significantly higher AL after adjustment. Hispanic/Latina participants did not have significantly elevated AL. Older age, greater area-level socioeconomic deprivation, and depression were independently associated with higher AL. Prior cancer diagnosis, cancer worry and perceived stress were not significantly associated with AL, whereas regular physical activity was associated with lower AL. Among cancer patients, advanced stage was associated with greater AL.

**Conclusions and Relevance:** This study demonstrates elevated AL among understudied racial/ethnic minority groups with familial cancer risk and identifies associations with neighborhood deprivation, depression, and physical activity. The association between cancer stage and AL suggests that physiologic stress may reflect variation in cancer burden. The lack of association with perceived stress and cancer worry further indicates that physiologic and self-reported psychosocial measures capture distinct dimensions of stress. The development of race/ethnicity-specific NLR thresholds derived from large population samples provide a benchmark for future studies.

## Introduction

Chronic stress is increasingly recognized as a contributor to health inequities and adverse cancer outcomes.^1–5^ Although stress affects all individuals, exposure to chronic psychosocial and environmental stressors is not equally distributed across populations.^6,7^ Racially and ethnically minoritized groups often experience disproportionate burdens of structural disadvantage, discrimination, economic insecurity, neighborhood deprivation, and barriers to health care, resulting in greater cumulative exposure to chronic stress across the life course. These exposures may contribute to persistent disparities in health outcomes through interconnected psychosocial and biologic pathways involving neuroendocrine activation, inflammation, metabolic dysregulation, and cardiovascular dysfunction.^6,8,9^

Allostatic load (AL) has emerged as a useful framework for quantifying the cumulative physiologic consequences of chronic stress exposure.^10–13^ AL reflects the cumulative “wear and tear” that occurs when physiologic stress-response systems are repeatedly activated over time.^6^ By integrating biomarkers across multiple physiologic systems, including cardiovascular, metabolic, inflammatory, and neuroendocrine pathways, AL provides a multidimensional composite index of biologic risk.^6,9,14–18^ Accordingly, quantitating AL may serve as an early biologic indicator of disease vulnerability and help identify potential modifiable targets for improved health outcomes.^7^

AL may represent the physiologic embodiment of social and structural inequity, providing a likely mechanistic link between chronic exposure to adverse conditions and downstream divergent health outcomes across racial/ethnic and socioeconomic groups.^19–22^ Growing evidence has demonstrated disproportionately higher AL among certain racially and ethnically minoritized populations, including Black and Hispanic/Latino, and among individuals experiencing socioeconomic disadvantage.^6,7,9,10,12,22^ Repeated exposure to social adversity may accelerate physiologic deterioration over time, through a process known as “weathering”, resulting in earlier biologic aging and greater disease burden among marginalized populations.^6,7^

However, direct comparisons of AL and its associations with psychologic stress, health outcomes and social determinants of health across diverse historically underrepresented minority (URM) populations using a common analytical framework remains limited.^9^

In this study, we examined psychological stress and AL among women enrolled in the Hereditary Exploration and Research for Screening and Testing for Oncology Risks in Women (HERSTORY) study, a cohort enriched for individuals from racial and ethnic groups that have been underrepresented in prior research and for families at increased risk for cancer. We evaluated differences in AL across diverse populations and introduced a novel AL metric incorporating population-specific neutrophil-to-lymphocyte ratio (NLR) thresholds. We also characterized the associations of AL with social determinants of health, psychosocial factors, health behaviors, and physiologic dysregulation among women who may experience both heightened cancer vulnerability and disproportionate exposure to social and structural stressors.

## Methods

### Study Population

We conducted a cross-sectional analysis of participants enrolled between October 2023 to September 2025 in the ongoing prospective UCLA HERSTORY cohort. HERSTORY was designed to evaluate genetic, clinical, environmental, and socioeconomic determinants of cancer risk among women with a family history of cancer and prioritizes recruitment of historically underrepresented racial and ethnic populations. Detailed race and ethnicity classifications are provided in the **Supplementary Methods**. Eligible participants were women aged ≥35 years, with a family history of cancer who provided informed consent. The study was approved by the UCLA Institutional Review Board (IRB-22-0771).

### Stress Measures by AL

Physiologic stress was quantified using AL. Because there is no universally accepted gold-standard definition of AL, biomarker selection was informed by prior literature and the availability of harmonized clinical laboratory data across cohorts.^18,23–26^ The full AL index included 13 measures spanning inflammatory, cardiometabolic, and organ-function domains: white blood cell count (WBC), absolute neutrophil count (ANC), NLR, BMI, hypertension, glucose or diabetes status, high-density lipoprotein cholesterol (HDL), total cholesterol-to-HDL ratio, triglycerides, cardiovascular morbidity, albumin, alkaline phosphatase, and estimated glomerular filtration rate.^27^

NLR varies across racial and ethnic groups making clinical interpretation and AL assessment difficult. We calculated race– and ethnicity-specific NLR tertiles derived using the most recent laboratory values from female patient in the UCLA Health population (n = 326,826).^28,29^ The derivation cohort, exclusions, group-specific sample sizes, and NLR thresholds are further described in the **Supplementary Methods.**

The complete scoring algorithm and thresholds are provided in **Table S1**. Because NLR, WBC, and ANC were missing in a subset of the HERSTORY cohort, the primary analysis used a 10-component AL score among participants with complete data (n = 303). High AL was defined as the upper tertile of the AL distribution within HERSTORY. Analyses comparing HERSTORY with matched ATLAS White participants used the 10-component score as the primary outcome and the full 13-component score in a secondary analysis using a subset of HERSTORY participants with all these components.

In HERSTORY, psychosocial stress was assessed using the 10-item Perceived Stress Scale (PSS-10; range, 0–40),^30,31^ and the 8-item Cancer Worry Scale (CWS-8; range, 8–32),^32^. High perceived stress was defined as a PSS-10 score ≥27, and high cancer worry as a CWS-8 score >14.^32,33^ Self-reported depression and prior cancer diagnoses were also captured.

Socioeconomic disadvantage was measured using the national Area Deprivation Index (ADI). Consistent with approaches used in prior studies, high neighborhood deprivation was defined as an ADI at or above the 80th percentile within HERSTORY.^34^

### HERSTORY and Matched ATLAS Cohorts

To contextualize the stress burden observed in the racially and ethnically diverse HERSTORY cohort, participants were matched with a reference cohort of self-reported non-Hispanic White females from the UCLA ATLAS biobank. The UCLA ATLAS Community Health Initiative enrolls participants across the UCLA Health system and reflects the broader Los Angeles population.^28^

Matched ATLAS cohort selection was designed to support HERSTORY comparisons across racial and ethnic groups and between participants with and without a prior cancer diagnosis. Detailed cohort construction is provided in the **Supplementary Methods** and **Table S2**.

For the primary analysis, we used a 10-component AL score. The final matched cohort included 709 participants: 468 non-Hispanic White participants from ATLAS and HERSTORY (the majority from ATLAS; with fewer than 10 from HERSTORY), and 241 HERSTORY participants who were not non-Hispanic White. Among HERSTORY participants, following filtering (see **Supplementary Methods**), 34 had a prior solid cancer diagnosis; these individuals were matched to 68 ATLAS participants with prior cancer. As a secondary analysis, we evaluated the consistency of the findings using the full 13-component AL score, incorporating inflammatory based metrics. This analysis included 359 participants: 125 from HERSTORY and 234 matched non-Hispanic White participants from ATLAS (see **Supplementary Methods).**

### HERSTORY and ATLAS Cancer-Only Cohorts

A cancer-focused analysis evaluated the association of AL (10-components) and cancer stage among participants with a solid cancer diagnosis before sample collection (see **Supplementary Methods**) and available information on stage, lab tests and BMI (n=227). The ATLAS cohort included 198 non-Hispanic White participants, and the HERSTORY cohort included 29 participants from racial and ethnic groups represented by more than five individuals. No matching was performed to preserve the available HERSTORY sample size; potential confounders were addressed through multivariable adjustment. Detailed distributions of stage and race/ethnicity are provided in the **Supplementary Methods**.

### Statistical Analysis

In the matched HERSTORY–ATLAS cohorts, multivariable linear regression evaluated associations of AL with race/ethnicity, age, ADI, prior cancer, and depression, and, in the cancer-only analysis, cancer stage and time since diagnosis. P values were adjusted using the false discovery rate (FDR) method, with an FDR-adjusted P value <0.10 considered statistically significant.

Within HERSTORY, participant characteristics and stress measures were summarized using descriptive statistics. Participants with and without a prior cancer diagnosis were compared using appropriate parametric or nonparametric tests (see the **Supplementary Methods** for more information).

## Results

### Racial and Clinical Determinants of Allostatic Load

The HERSTORY cohort included 303 female participants with a median age of 55 years (IQR, 46–64). The cohort was intentionally racially and ethnically diverse; 38.9% of participants were Black, 23.1% were Hispanic/Latina, and 22.1% were Asian or Pacific Islander; smaller proportions identified as multiracial, American Indian/Alaska Native, non-Hispanic White, or Caribbean/West Indian (**Table 1**).

**Table 1.** Baseline Demographic characteristics of participants in the HERSTORY cohort at study enrollment (N = 303)

| Variable | Cohort<br>N = 303 |
| --- | --- |
| **Demographics** |  |
| **Age** |  |
| Mean (SD) | 55.6 (11.86) |
| Median (IQR) | 55 (46, 64) |
| **Race/Ethnicity, n** |  |
| AIAN | <10 |
| Asian/Pacific Islander | 67 |
| Black | 118 |
| Hispanic/Latina | 70 |
| Non-Hispanic White | <10 |
| Multiracial | 37 |
| Caribbean/West Indian | <10 |
| **Clinical Factors** |  |
| **Past cancer diagnosis, n (%)** |  |
| Yes | 45 (14.9) |
| No | 258 (85.1) |
| **BMI, n (%)** |  |
| Median (IQR) | 27.3 (24.0-31.3) |
| Underweight/Normal | 102 (33.7) |
| Overweight | 104 (34.3) |
| Obese | 97 (32.0) |
| **Hypertension, n (%)** | 106 (35.0) |
| **Diabetes mellitus, n (%)** | 35 (11.6) |
| **Hypercholesterolemia, n (%)** | 99 (32.7) |
| **Exercise in past 12 months (moderate–vigorous physical activity), n (%)** |  |
| None | 28 (8.6) |
| ≤ 1 day per week | 114 (38.0) |
| 2-3 days per week | 93 (30.7) |
| 4-7 days per week | 70 (23.1) |
| Missing |  |

**Acronyms:** AIAN- American Indian or Alaska Native; BMI-body mass index

**Table 2.** HERSTORY Psychosocial Stress Measures, Cancer History, and Social Determinants of Health stratified by Allostatic Load (n=303)

|  | Low AL<br>N = 127 | Moderate AL<br>N = 87 | High AL<br>N = 89 |
| --- | --- | --- | --- |
| **Psychosocial stress measures** |  |  |  |
| **Perceived Stress Scale (PSS-10)** |  |  |  |
| Mean (SD) | 14.5 (6.84) | 15.1 (6.36) | 14.3 (7.86) |
| Low ($\leq 13$), n (%) | 65 (51.2) | 32 (36.8) | 45 (50.6) |
| Moderate (14-16) or High ($\geq 27$), n (%) | 62 (48.8) | 55 (63.2) | 44 (49.4) |
| **Cancer Worry Scale (CWS-8)** |  |  |  |
| Mean (SD) | 14.1 (4.22) | 13.9 (3.77) | 14.3 (4.62) |
| High ($> 14$), n (%) | 49 (38.6) | 35 (40.2) | 33 (37.5) |
| **Depression**, n (%) | 21 (16.5) | 19 (21.8) | 25 (28.1) |
| **Past cancer diagnosis**, n (%) | 17 (13.4) | 11 (12.6) | 17 (19.1) |
| **Social Determinants of Health** |  |  |  |
| **State Area Deprivation Index (ADI)** |  |  |  |
| Median (IQR) | 3 (2, 4) | 3 (2, 4) | 3 (2, 6) |
| **National Area Deprivation Index (ADI)** |  |  |  |
| Median (IQR) | 5.5 (3, 11) | 7 (3, 12) | 7 (3, 15) |

Values are mean (SD) unless otherwise specified
Stress measures by AL tertile – only among patients with no missing values across subset of 10 components (N = 303)

To evaluate racial and ethnic differences in AL, we compared HERSTORY participants with matched non-Hispanic White female participants from ATLAS, serving the same Los Angeles geographic region (n=709; **Figure 1a**).^28^ AL was first calculated using 10 commonly used cardiovascular and metabolic markers (**Figure 1b**). Mean AL scores were 2.1 (SD, 1.5) in matched HERSTORY participants and 2.0 (SD, 1.7) in ATLAS participants. Among HERSTORY participants, 14.9% had high AL, defined as the highest HERSTORY tertile and corresponding to a score >2.5 (**Table S3**).

**Figure 1.**
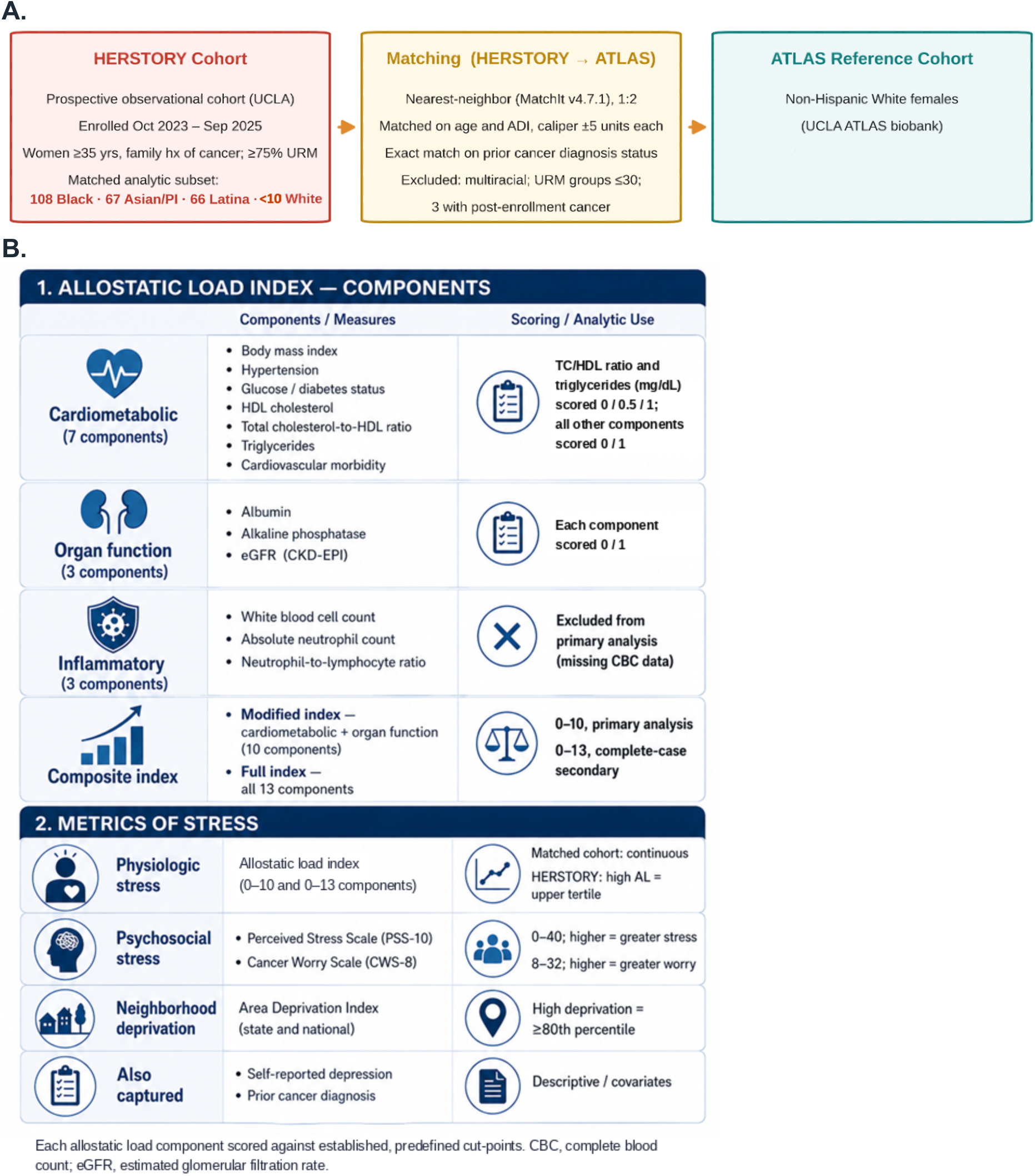
**A**. HERSTORY and ATLAS Matched cohort Study and **B.** Allostatic Load and Stress Metrics

After adjustment for age, national Area Deprivation Index (ADI), prior cancer diagnosis, and depression, Black participants (β = 0.37; FDR = 0.04) and Asian or Pacific Islander participants (β = 0.35; FDR = 0.09) had higher AL than non-Hispanic White participants. AL was not significantly elevated among Hispanic/Latina participants (β = 0.25; FDR = 0.22; **Table 3A; Figure 2a; Figure S1a**). These findings confirm higher AL in Black individuals and reveal previously unreported elevated AL in Asian or Pacific Islander individuals relative to White participants.^35–38^

**Figure 2.**
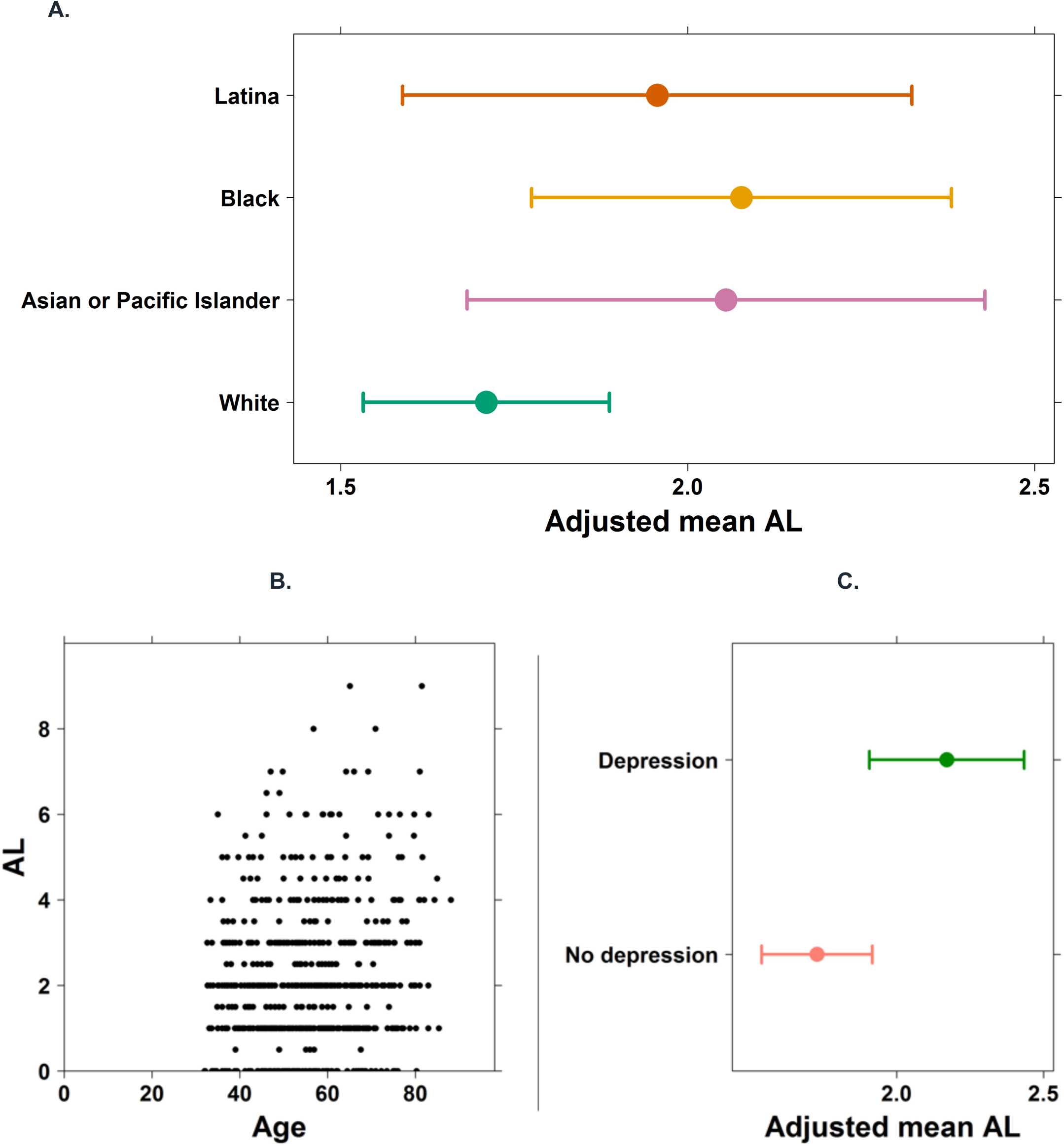
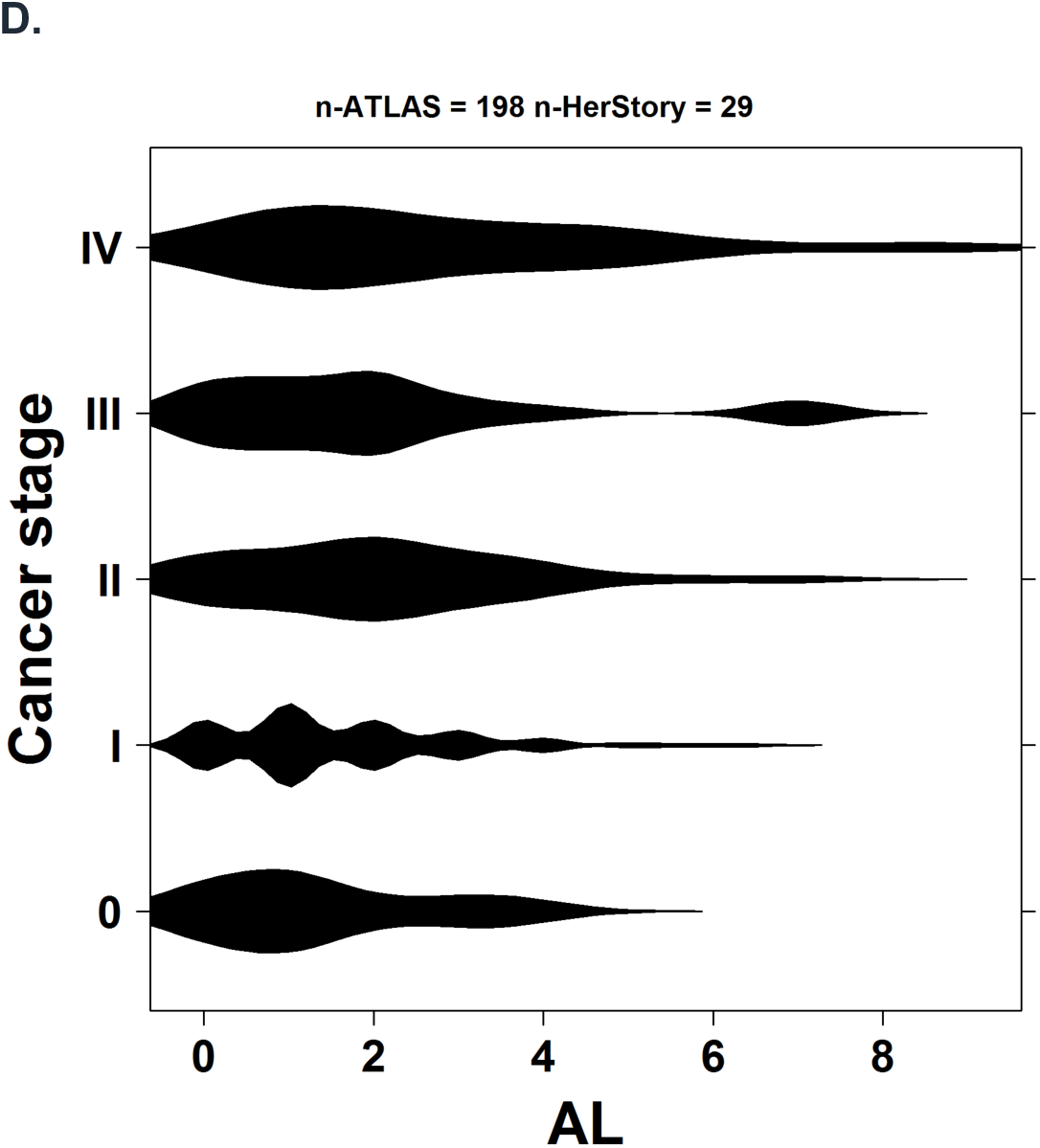
Associations between Allostatic Load (score 0-10) and **(A)** Race/Ethnicity, **(B)** Age, and **(C)** Depression are shown among participants in the HERSTORY and ATLAS Matched cohort. Figures A and C show adjusted means. (n=709). **(D)** Associations between Allostatic Load (score 0-10) and Cancer Stage in HERSTORY and ATLAS Pooled cohort (n=227). For categorical predictors (panels A and C), adjusted means estimated using the emmeans function^65^ are presented for race (A) and depression (C) categories. These estimates (A-C) account for other covariates in the model, providing a more representative assessment of differences between groups.

**Table 3A.** Multivariable Model of Allostatic Load in the HERSTORY Underrepresented Minority Population and Matched Non-Hispanic White ATLAS Cohort 10-component Allostatic Load (n=709)

| Variable | $\beta$ estimate | Standard error | FDR-adjusted P value |
| --- | --- | --- | --- |
| Previous cancer diagnosis | 0.017 | 0.157 | 0.914 |
| **Asian or Pacific Islander vs non-Hispanic White** | **0.345** | **0.188** | **0.093** |
| **Black vs non-Hispanic White** | **0.368** | **0.153** | **0.039** |
| Latina vs non-Hispanic White | 0.246 | 0.187 | 0.220 |
| **Age at laboratory testing, per year** | **0.024** | **0.005** | **<0.001** |
| **National Area Deprivation Index, per unit** | **0.015** | **0.007** | **0.049** |
| **Depression** | **0.442** | **0.124** | **0.001** |

Note: AL excluded NLR, ANC, and WBC
Bold values indicate FDR-adjusted $P < 0.10$ . $\beta$ , regression coefficient;
Abbreviations: AL-allostatic load; FDR-false discovery rate; NLR-neutrophil-to-lymphocyte ratio; WBC-white blood cell count; ANC-absolute neutrophil count

**Table 3B.**
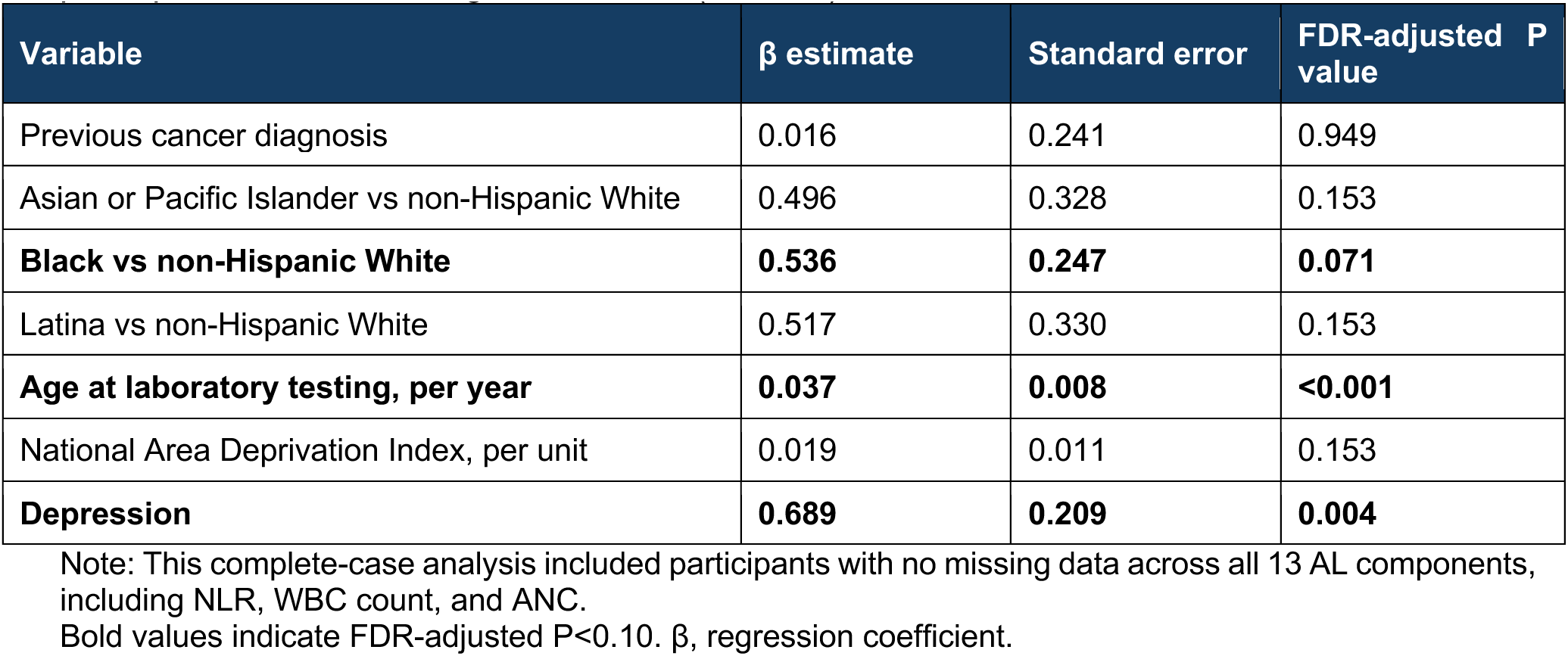
Multivariable Model of Allostatic Load in the HERSTORY Underrepresented Minority Population and Matched Non-Hispanic White ATLAS 13-component Allostatic Load among participants without missing AL variables (N = 359)

| Variable | $\beta$ estimate | Standard error | FDR-adjusted P value |
| --- | --- | --- | --- |
| Previous cancer diagnosis | 0.016 | 0.241 | 0.949 |
| Asian or Pacific Islander vs non-Hispanic White | 0.496 | 0.328 | 0.153 |
| **Black vs non-Hispanic White** | **0.536** | **0.247** | **0.071** |
| Latina vs non-Hispanic White | 0.517 | 0.330 | 0.153 |
| **Age at laboratory testing, per year** | **0.037** | **0.008** | **<0.001** |
| National Area Deprivation Index, per unit | 0.019 | 0.011 | 0.153 |
| **Depression** | **0.689** | **0.209** | **0.004** |

Note: This complete-case analysis included participants with no missing data across all 13 AL components, including NLR, WBC count, and ANC.
Bold values indicate FDR-adjusted $P < 0.10$ . $\beta$ , regression coefficient.

As already established in prior literature, increasing age (β = 0.02; FDR = <0.01), depression (β = 0.44; FDR <0.01), and greater neighborhood deprivation (β = 0.02; FDR = 0.05) were also associated with higher AL (**Figure 2b–c; Figure S1b**).^39–41^

We next examined whether incorporating less commonly used inflammatory hematologic markers altered these findings. A 13-component AL score was constructed by adding NLR, ANC, and WBC to the original measure. To account for population variation in NLR, race-– and ethnicity-stratified tertiles were derived from the entire UCLA Health female population (n = 326,826),^28,29^ which, to our knowledge, is the largest population used to establish race– and ethnicity-specific NLR thresholds.

Because complete inflammatory-marker data was available for fewer participants in HERSTORY, this analysis included a reduced pooled sample with complete data (n=359). Higher AL among Black participants remained significant, and the association among Asian or Pacific Islander participants remained directionally consistent but was no longer statistically significant, likely reflecting the approximately 50% reduction in sample size. The AL associations with age and depression remained significant (**Table 3B**).

Although the comparative performance of the 10– and 13-component measures could not be directly compared because they were evaluated in different samples, these findings suggest that these inflammatory measures and novel population-based NLR thresholds are compatible with established AL framework.

### HERSTORY Psychosocial Stress, Depression, and Neighborhood Deprivation

Among HERSTORY participants, the mean perceived stress score was 14.6 (SD, 7.01): 46.9% reported low stress, 47.2% moderate stress, and 5.9% high stress. The mean cancer worry score was 14.1 (SD, 4.21), with 38.6% classified as having high cancer worry. Depression was self-reported by 21.5% of participants.

Median national ADI was 6 (IQR, 3–12). Perceived stress and cancer worry did not differ significantly across AL groups (low, n = 127; moderate, n = 87; high, n = 89), suggesting that contemporaneous self-reported stress was not closely aligned with cumulative physiologic stress burden (**Table 2**).

Consistent with the matched pooled analysis, both depression and neighborhood deprivation were independently associated with high AL in adjusted analyses (**Table S4**). Participants with a national ADI ≥15 had more than threefold higher odds of high AL than those living in less deprived areas (adjusted OR, 3.32; 95% CI, 1.59–6.93), while participants with depression had nearly threefold higher odds (adjusted OR, 2.74; 95% CI, 1.28–5.86).

### AL and Prior Cancer Diagnosis

A prior cancer diagnosis was reported by 14.9% of HERSTORY participants (n=45). Prior cancer diagnosis was not significantly associated with AL in HERSTORY alone (**Table S5)** or in the pooled HERSTORY–ATLAS analysis after adjustment (β = 0.017; FDR = 0.914). However, in a cancer focused analysis cancer stage was positively associated with AL using both the 10-component score (β = 0.30; FDR = 0.003; **Figure 2d**) and even more strongly when inflammatory components were integrated (13-component score; β = 0.40; FDR = 0.0006; **Figure S1c**). AL was not associated with cancer-related pathogenic germline variant status.

### Clinical Components of AL and Clinical Characteristics

To better characterize the determinants of AL in URM populations, we examined the individual clinical and behavioral factors contributing to the AL index among HERSTORY participants (n = 303). Among participants with complete data for the 10-component AL measure, the most common adverse component was BMI outside the normal range (67.7%; median BMI, 27.3), followed by hypertension (35.0%) and elevated total cholesterol-to-HDL cholesterol ratio (28.3) (**Table S3**). Exercising at least 4 days per week was independently associated with lower odds of high AL (adjusted OR, 0.32; 95% CI, 0.12–0.89).

## Discussion

In this racially and ethnically diverse cohort of women with familial cancer risk, Black and Asian or Pacific Islander participants had higher AL than matched non-Hispanic White participants. The higher AL observed in Black participants is consistent with previous research, whereas the association among Asian or Pacific Islander participants addresses an important gap. Large U.S. studies have primarily examined Black-White and Hispanic/Latino-White disparities, while Asian Americans, the fastest-growing racial group in the U.S., have frequently been excluded, or classified as “other”, rendering their experiences largely invisible.^42–44^ Our findings underscore the importance of adequately sampling these heterogeneous populations, particularly in diverse metropolitan regions such as Los Angeles County, where Asian populations represent approximately 16.6%.^45^

Hispanic/Latina participants, who represented a large proportion of HERSTORY and 49.3% of Los Angeles county, did not have significantly higher AL than matched non-Hispanic White participants, differing from prior studies.^46–48^ Associations between Hispanic/Latino identity and AL may vary according to nativity, acculturation, socioeconomic context, family and community support and other sources of resilience.^47,49,50^ The large and established Hispanic/Latino Los Angeles County population may provide social networks that buffer chronic stress. Nevertheless, the association between neighborhood deprivation and higher AL highlights the contribution of structural and socioeconomic conditions to cumulative physiologic burden.

Increasing age was strongly associated with AL, consistent with its conceptualization as cumulative multisystem wear and a marker of accelerated biological aging.^51–54^ Previous studies have linked elevated AL with depression among patients with cancer, particularly those with lower income or greater exposure to structural adversity and our results reinforce the close relationship.^55–59^ Depression may contribute to physiologic strain through neuroendocrine dysregulation and systemic pro-inflammatory signaling, creating a cycle in which structural adversity contributes to depression and chronic stress signaling further promotes cardiometabolic and inflammatory dysregulation captured by AL. These findings support integrating mental health assessment and treatment into care for populations at increased familial cancer risk, particularly those exposed to a greater social and structural stressors.

Psychosocial adaptation and physiologic stress burden represent distinct but partially overlapping dimensions of stress. Subjective psychological resilience and adaptation, as measured by self-reported stress and cancer worry, may coexist with unrecognized systemic physiologic burden, especially in individuals experiencing depression or socioeconomic deprivation.

Patient-reported measures remain essential for understanding perceived emotional well-being, coping, and cancer-related concerns, but they may not fully capture multisystem dysregulation. This distinction may be especially relevant in populations exposed to social, economic, or healthcare-related adversity without reporting high contemporaneous stress. Therefore, AL may complement psychosocial screening by identifying individuals with elevated physiologic risk who might otherwise appear psychologically well adapted.

The protective association between regular physical activity and AL in the HERSTORY cohort is clinically meaningful. Although causality cannot be inferred from this cross-sectional study, the finding is consistent with evidence that lifestyle and behavioral interventions may mitigate physiologic stress burden.^60^ Together with the associations observed for depression and neighborhood deprivation, these results suggest that AL is not simply a fixed index of prior adversity but may identify potentially modifiable targets for intervention. Exercise, cardiometabolic risk management, depression treatment, and community-based stress-reduction programs may therefore represent practical strategies for reducing physiologic burden in high-risk populations.^61,62^

AL was not driven by hereditary cancer susceptibility or a history of cancer. These findings suggest that social, behavioral, and mental health factors may contribute more strongly to physiologic stress burden than genetic cancer predisposition itself. However, the modest number of participants with prior cancer limits definitive conclusions. To our knowledge, this is the first study to identify an association between cancer stage and AL. Given the limited sample size of underrepresented minority individuals in the cancer-focused analyses, larger studies are needed to better characterize this association within these populations.

This study has several strengths. It evaluates AL in a racially and ethnically diverse cohort enriched for populations historically underrepresented in stress-biology research and integrates physiologic, psychosocial, behavioral, and neighborhood-level measures. It also evaluates the addition of inflammatory hematologic markers and establishes race– and ethnicity-specific NLR thresholds using a clinical population approximately 35 times larger than the largest previously reported mixed-population cohort of 9,427 US adults.^63,64^ These thresholds may provide a useful benchmark for future studies.

Despite these strengths, several limitations should be considered. The cross-sectional design precludes causal inference and prevents evaluation of temporal relationships between AL, psychosocial stress, cancer, and depression. Depression and some AL components were ascertained differently in HERSTORY and ATLAS. Depression was self-reported in HERSTORY but identified using diagnostic codes in ATLAS, potentially underestimating its prevalence in both cohorts. ATLAS participants were recruited through a health-system biobank and may differ from HERSTORY participants in healthcare engagement, referral patterns, comorbidity burden, and the clinical context of laboratory testing. Matching and multivariable adjustment were used to mitigate these differences, and laboratory measurements or cancer diagnoses were required to precede ATLAS enrollment.

The inflammatory-marker analysis was limited to a smaller complete-case sample. In addition, small subgroup sizes prevented disaggregation of Hispanic/Latina participants and Asian and Pacific Islander participants into more specific populations. Such aggregation may obscure meaningful heterogeneity in social exposures, health behaviors, and physiologic risk. Larger studies should oversample and disaggregate these populations whenever feasible.

## Conclusion

This study identifies racial and ethnic differences in cumulative physiologic stress burden and supports a multidimensional approach to familial cancer risk care that integrates addressing patient-reported stress, mental health, physiologic burden, and social determinants of health. Longitudinal studies are needed to determine whether behavioral, mental health, and structural interventions can reduce AL and improve long-term health outcomes in historically underserved populations.

## Supplementary Methods

### HERSTORY Cohort

The HERSTORY study integrates sociodemographic data, psychosocial assessments, clinical information, and biospecimen collection, providing a unique opportunity to evaluate both perceived and physiologic stress burden among women with familial cancer risk, including those with and without a personal history of cancer.

Exclusion criteria were age <35 years, male sex assigned at birth, absence of a family history of cancer, prior bone marrow transplantation, or inability or unwillingness to provide informed consent.

Participants were required to provide informed consent, authorize access to clinical data, and undergo clinical genetic testing, including whole-genome sequencing if testing had not previously been completed.

### Study Race and Ethnicity Classification

Race and ethnicity were categorized using participant-reported responses. Participants identifying as Hispanic or Latina were classified as Hispanic/Latina regardless of reported race, including those who also identified as White. The White category was restricted to participants who identified as White and reported non-Hispanic/Latina ethnicity or declined to report ethnicity. Participants selecting “Other” were reclassified using their accompanying race or ethnicity description: those reporting Hispanic/Latina ethnicity were categorized as Hispanic/Latina; those identifying as White, Middle Eastern, or Afro-Caribbean were categorized as White, Middle Eastern, or Caribbean/West Indian, respectively; and participants reporting other combinations of racial or ethnic identities were categorized as mixed race/ethnicity. Participants selecting “Other” who reported non-Hispanic/Latina ethnicity or declined to report ethnicity and provided no further classifiable information were retained in the Other category.

### NLR Benchmarks Across Racial and Ethnic Populations

While ATLAS is a biobank comprising a subset of UCLA Health patients who consented to linked genetic and EHR research, the UCLA Health population includes all patients receiving care within the health system and does not include linked genetic data.^28,29^ This population was substantially larger than that included in the previous largest analysis, which used National Health and Nutrition Examination Survey data from 9,427 U.S. adults enrolled between 2007 and 2010 to characterize NLR by race and ethnicity.^63^ The number of individuals used to obtain NLR tertiles for every race group were: 198,607 White non-Hispanic/Latina, 29,996 White Hispanic/Latina, 56,257 Asian, 27,306 Black, 11,432 Middle Eastern or North African, 1,455 American Indian/Alaska Native, 1,005 Pacific Islander, 718 Caribbean/ West Indian. Exclusion criteria included individuals with unknown race, non-White Hispanic/Latina, other smaller race groups, male participants, and individuals with neutrophil or lymphocyte counts of zero.

Given the small number of Pacific Islander participants in HERSTORY (n<10), Asian and Pacific Islander individuals were combined in the primary analyses to maintain adequate statistical power. Nevertheless, the NLR threshold incorporated into the AL score was specified separately for Pacific Islander participants, reflecting potential physiologic heterogeneity within this aggregated racial and ethnic category.

### HERSTORY and Matched ATLAS Cohorts

HERSTORY participants were compared with self-reported non-Hispanic White female participants from the UCLA ATLAS biobank. ATLAS participants laboratory and body mass index (BMI) values were selected relative to the cancer diagnosis and laboratory dates using prespecified time windows. Laboratory values for this group were selected as close as possible to the cancer diagnosis date with the maximum interval of 14 years, corresponding to the longest interval observed in the HERSTORY cohort. BMI values were assigned using the measurement closest to the laboratory date, allowing BMI values obtained up to one month after laboratory testing.

Comorbidities were identified using Classification of Diseases (ICD) codes (**Table S3**). ATLAS laboratory values used to calculate AL were restricted to measurements obtained before biospecimen collection.^66^ For ATLAS participants with cancer, solid tumor diagnoses were identified using ICD codes, and were required to precede biospecimen collection.

Among participants with cancer, diagnoses of depression, hypertension, cardiovascular disease, and diabetes were defined based on codes recorded between the cancer diagnosis date and the laboratory test date. Among participants without cancer, these conditions were defined based on codes recorded before the laboratory testing date.

The final matched cohort comprised 709 participants: 468 non-Hispanic White participants from ATLAS and HERSTORY (the majority from ATLAS; HERSTORY < 10), and 241 HERSTORY participants, comprising 67 Asian or Pacific Islander, 108 Black, and 66 Hispanic/Latina participants. Among HERSTORY participants, 34 had a prior solid cancer diagnosis. These individuals were matched to 68 ATLAS participants with prior cancer.

For matching, HERSTORY participants from non-White racial and ethnic groups represented by more than 30 individuals were included, along with the small number of non-Hispanic White participants; participants who identified as multiracial were excluded. HERSTORY participants who developed cancer after enrollment were also excluded. Nearest-neighbor matching was performed using the MatchIt R package, (version 4.7.1)^67^, matching participants on age and ADI using a ±5-point caliper in a 1:2 ratio, with exact matching required for cancer diagnosis status. Due to the ADI matching criteria, nine HERSTORY individuals were excluded from the analysis, and 33 individuals had only one available ATLAS match rather than two. Covariate balance after matching was assessed using standardized mean differences (SMDs). All covariates achieved an absolute SMD < 0.1 after matching (age 0.032 and ADI 0.099), indicating good covariate balance. A more permissive ADI caliper was not used because it resulted in poorer covariate balance.

To examine the consistency of AL index with the integration of the inflammatory hematologic measures, we evaluated a complete-case analysis included 359 participants with no missing components across all 13 AL variables: 234 matched non-Hispanic White ATLAS and HERSTORY participants (the majority from ATLAS, with fewer than 10 from HERSTORY), and 122 additional HERSTORY participants (61 Black, 31 Asian or Pacific Islander, 30 Hispanic/Latina)

### HERSTORY and ATLAS Cancer-Only Cohorts

A secondary cancer-only analysis was conducted among participants with a prior solid cancer diagnosis and available cancer stage information. The ATLAS comparison cohort included self-reported non-Hispanic White female participants with a solid cancer diagnosis and documented cancer stage. Consistent with the primary analysis, only cancers diagnosed before biospecimen collection were included. The ATLAS cancer-only cohort comprised participants with stage 0 (n = 14), stage I (n = 106), stage II (n = 28), stage III (n = 18), and stage IV disease (n = 32).

Within HERSTORY, 29 participants had a prior solid cancer diagnosis and available stage information. To support meaningful subgroup comparisons, only racial and ethnic groups with more than five participants were included (Asian or Pacific Islander, Black, and Hispanic/Latina participants). The corresponding stage distribution was stage 0 – I (n = 19) and stage > II (n = 10) (exact stage was used in the analysis). No matching was performed in the cancer-only analysis to preserve the available sample size. Potential confounding variables were instead accounted for through statistical adjustment.

### Statistical Analysis

Within HERSTORY, descriptive statistics were used to summarize baseline characteristics and stress measures. Comparisons between HERSTORY participants with and without prior cancer were performed using appropriate parametric or nonparametric tests. Multivariable logistic regression models were developed to identify predictors of high AL, defined as the upper tertile within the HERSTORY cohort. To improve interpretability and reduce the influence of sparse categories, several continuous and ordinal predictors were dichotomized prior to modeling, including age (>55 years vs. ≤55 years), national ADI rank at the 80^th^ percentile within the HERSTORY cohort (≥15 vs. 0–14), and physical activity (≥4 days per week of moderate-to-vigorous activity versus less than 4 days). Perceived stress and cancer worry were each collapsed into binary high/low categories. Prior cancer diagnosis and self-reported depression were also included. Unadjusted and adjusted odds ratios with 95% confidence intervals were reported for each predictor.

In the matched HERSTORY–ATLAS cohorts, multivariable linear regression evaluated associations of AL scores. Models included race/ethnicity, age, ADI, prior cancer diagnosis, and depression. For the analysis of HERSTORY and ATLAS cancer-only cohorts, models included cancer stage, and years from cancer diagnosis to AL measurement, in addition to above predictors. Non-Hispanic White race/ethnicity, no prior cancer diagnosis, and no depression served as reference categories. To account for multiple comparisons, P values were adjusted using the false discovery rate (FDR) method, with an FDR-adjusted P value <0.10 considered statistically significant. Non-Hispanic White race/ethnicity and\ no depression were the reference categories.

Figures involving HERSTORY and ATLAS participants were generated using the Boutros Plotting General R package (version 7.1.2).^65^ Estimated marginal means were calculated using the emmeans package (version 1.8.8).^68^

## Data Sharing Statement

Deidentified HERSTORY participant data used in the analyses may be made available from the corresponding author upon reasonable request and approval by the study investigators and relevant institutional authorities. The shared materials may include the deidentified analytic data set and accompanying data dictionary. Data will be provided for approved research purposes, subject to applicable institutional policies, data use agreements, and protections for participant confidentiality. Access to ATLAS data may be requested through collaboration.

## Acknowledgment and Funding Support

The authors would like to thank April Krueger for the residual class settlement funds in the matter of April Krueger v. Wyeth, Inc., Case No. 03-cv-2496 (US District Court, SD of Calif.) that funded this study. We are deeply grateful for each of the HERSTORY participants for generously contributing their time and effort to advance this work.

We gratefully acknowledge the support of the Institute for Precision Health, participating patients from the UCLA ATLAS Precision Health Biobank, UCLA David Geffen School of Medicine, UCLA Clinical and Translational Science Institute (CTSI) grant number UL1TR001881, and UCLA Health. We thank the Biomedical Informatics Program at UCLA CTSI and the Office of Health Informatics and Analytics for providing access to the Data Discovery Repository.

Eliya K. Shachar and Victoria E. Rodriguez were supported by the National Institutes of Health, National Cancer Institute T32 Patient-Centered Outcomes Research Training in Urologic and Gynecologic Cancers Program (T32CA251072). Roni Haas was supported by the Prostate Cancer Foundation Young Investigator Award (22YOUN32).

